# The role of artificial intelligence in the application of the integrated electronic health records and patient-generated health data

**DOI:** 10.1101/2024.05.01.24306690

**Authors:** Jiancheng Ye, Jiarui Hai, Jiacheng Song, Zidan Wang

## Abstract

**Objective:** This scoping review aims to identify and understand the role of artificial intelligence in the application of integrated electronic health records (EHRs) and patient-generated health data (PGHD) in health care, including clinical decision support, health care quality, and patient safety. We focused on the integrated data that combined PGHD and EHR data, and we investigated the role of artificial intelligence (AI) in the application in health care.

**Methods:** We used the Preferred Reporting Items for Systematic Reviews and Meta-Analyses (PRISMA) guidelines to search articles in six databases: PubMed, Embase, Web of Science, Scopus, ACM Digital Library, and IEEE Computer Society Digital Library. In addition, we synthesized seminal sources, including other systematic reviews, reports, and white papers, to inform the context, history, and development of this interdisciplinary research field.

**Results:** Fifty-six publications met the review criteria after screening. The EHR-integrated PGHD introduces benefits to health care, including empowering patients and families to engage via shared decision-making, improving the patient-provider relationship, and reducing the time and cost of clinical visits. AI’s roles include cleaning and management of heterogeneous datasets, assisting in identifying dynamic patterns to improve clinical care processes, and providing more sophisticated algorithms to better predict outcomes and propose precise recommendations based on the integrated data. Challenges mainly stem from the large volume of integrated data, data standards, data exchange and interoperability, security and privacy, interpretation, and meaningful use.

**Conclusion:** The use of PGHD in health care is at a promising stage but needs further work for widespread adoption and seamless integration into health care systems. AI-driven, EHR-integrated PGHD systems can greatly improve clinicians’ abilities to diagnose patients’ health issues, classify risks at the patient level by drawing on the power of integrated data, and provide much-needed support to clinics and hospitals. With EHR-integrated PGHD, AI can help transform health care by improving diagnosis, treatment, and the delivery of clinical care, thus improving clinical decision support, health care quality, and patient safety.

## INTRODUCTION

The recent widespread routine uses of smartphones, wearable sensors, and the Internet of Things (IoT) technologies have emerged as promising data sources for health care research.[1] These technologies are driving more targeted patient engagement thanks to the massive development of digital capacities. Personal health information has also spiked with the broad availability of low-cost devices designed to monitor and inform healthy lifestyles.[2] This scoping review focuses on the intersection of artificial intelligence (AI), electronic health records (EHRs), and patient-generated health data (PGHD) in health care, including clinical decision support, health care quality, and patient safety.

### Patient-generated health data

Patient-generated health data (PGHD), defined by the Office of the National Coordinator for Health Information Technology (ONC), includes health symptoms, medical history, biometric information, treatment history, lifestyle data, and other information.[3] PGHD is created, gathered, and recorded either by patients or their designees (i.e., care partners or those who assist them) to help address a health concern.[4] By reviewing and discussing PGHD with patients remotely, clinicians can address clinical issues efficiently outside of clinical settings.

PGHD allows health issues to be closely monitored without inconvenient traffic, insurance copayments, or lost days at work. It also provides actionable information for disease risk assessment and identification of issues that require urgent attention. In addition, PGHD can improve provider-patient communication and relationships by promoting mutual understanding, follow-up and feedback,[5, 6] and thought sharing.[7] Various opportunities regarding PGHD are envisioned in the context of big data and artificial intelligence (AI) in health care. The ONC has initiated several activities to advance knowledge of the PGHD and identify policies and promising practices to support this field.

### Artificial intelligence

AI is a technology that aims to mimic human cognitive functions in the analysis, presentation, and comprehension of complex health data and bring a revolutionary paradigm innovation to health care. This innovation is powered by the increased availability of health data (structured and unstructured) and rapid progress of analytical techniques. AI has unique abilities to collect and gather data, process it, and give a well-defined output to the end-user. AI can use sophisticated algorithms to learn features from a large volume of health-related data, and then use the obtained insights to assist clinical practice. Embedding AI technologies into health care can help to reduce diagnostic and therapeutic errors that are inevitable in human practice, thus improving health care quality and patient safety. AI can also extract useful information from a large patient population data, including PGHD and EHR, to assist in making real-time inferences for health outcome prediction and health risk alerts. AI technologies have been successfully applied to clinical practices such as diagnosis processes,[8] treatment protocol development,[9] personalized medicine,[10] drug development,[11] and patient monitoring and care.[12] Various specialties in health care have shown an increase in research related to AI.[13] Currently, many countries, including the United States, are investing billions of dollars to support the development of AI in health care.[14].

### Clinical decision support

Clinical decision support (CDS) systems provide health care workers with essential knowledge, intelligently manage electronic health data, present critical information at appropriate times, produce clinical advice, and thus enhance health care. CDS encompasses a variety of tools to enhance decision making in the clinical workflow, including clinical guidelines, documentation templates, computerized alerts and reminders to clinicians and patients, patient reports and summaries, diagnostic support, and contextually relevant reference information.[15]

The Institute of Medicine (IOM) also promotes the use of CDS systems to advance the quality of patient care.[16] CDS has contributed to health care by minimizing medical errors (e.g., medication prescription errors, adverse drug events, and administration errors), providing tools to measure clinician performance and patient outcomes,[17] and simplifying the workflow by integrating real-time PGHD into EHR.

### Health care quality

The Institute of Medicine (IOM) defines quality of care as “the degree to which health services for individuals and populations increase the likelihood of desired health outcomes and are consistent with current professional knowledge”.[18] The World Health Organization (WHO) states that health care quality should have three characteristics: effectiveness, safe, and people-centered.[19] Examples include therapy’s reduction or lessening of diseases identified by medical diagnosis, decrease in the number of risk factors, or health indicators in the population who are accessing certain kinds of care. Health care quality improvement began in the 19^th^ century, and interventions were implemented in an effort to improve health care outcomes.[20, 21] More recently, a focus on health care quality improvement has been the emerging health information technologies (e.g., EHR, PGHD).[22]

### Patient safety

Patient safety is defined as the minimization of the risk of harm associated with the health care process and the absence of preventable harm to patients.[23] Patient safety aims to protect patients through the prevention, reduction, reporting, and analysis of errors and other types of unnecessary harm that often lead to adverse events.[24] In 1999, the IOM released the report, “To Err is Human: Building a Safer Health System,” which called for a broad effort to establish a Center for Patient Safety, develop safety programs in health care organizations, expand reporting on adverse events, and further engage policymakers, health care workers, professional societies, and researchers in achieving these goals .[25] Despite these post-IOM report initiatives, studies have shown that medical errors are still the third leading cause of death in the US, and over 250,000 people die every year due to medical mix-ups.[26]

Integrating PGHD into EHRs further expands the capacity to monitor patients’ health status without requiring office visits or hospitalizations. The combination of PGHD and EHR enables health care providers to have a more comprehensive view of the patients. Although there is increasing attention on the application of AI using either EHR or PGHD (**Figure 1a** and **1b**), little is known about its application to integrated EHR and PGHD data (**Figure 1c**). It would be valuable to (1) investigate and understand the current state of the integration of PGHD into EHR with a focus on AI application in clinical care, and (2) identify potential roles of AI in the efficient translation of EHR-integrated PGHD into health care applications.[27]

**Figure 1.**
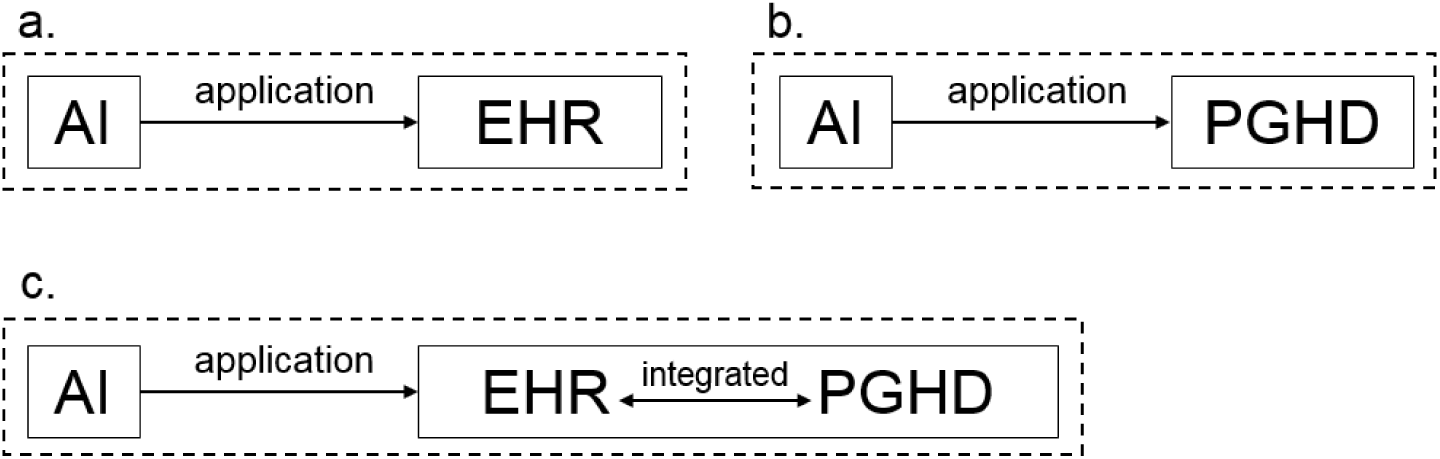
The association of artificial intelligence applications with EHR, PGHD, and EHR-integrated patient generated health data. AI= artificial intelligence, EHR= electronic health records, PGHD= patient-generated health data.

This scoping review aims to identify and understand the role of AI in the application of the EHR-integrated PGHD in health care, including clinical decision support, health care quality, and patient safety (**Figure 2**). We focus on applications of AI that use the integrated data and identify opportunities, successful experiences, challenges, strategies, and implications.

**Figure 2.**
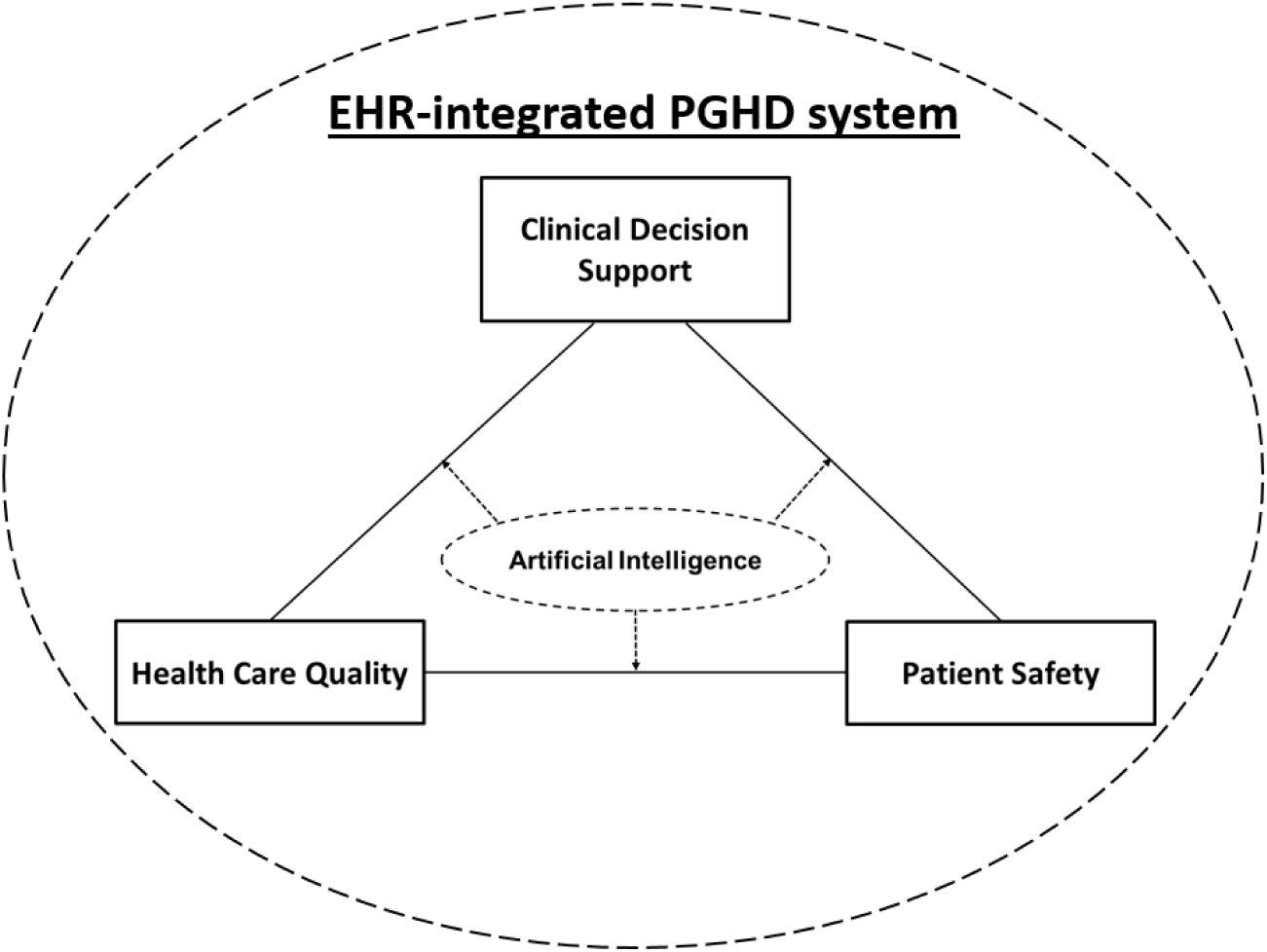
The framework of artificial intelligence application of electronic health record (EHR)–integrated patient-generated health data (PGHD) system on clinical decision support, health care quality, and patient safety.

## MATERIALS AND METHODS

We designed and reported this scoping review according to the Preferred Reporting Items for Systematic Reviews and Meta-Analyses (PRISMA).[28] In addition, we also synthesized seminal sources, including other systematic reviews, reports, and white papers, to inform the context, history, and development of this interdisciplinary research field.

### Search strategy

We searched six databases: PubMed, Embase, Web of Science, Scopus, ACM Digital Library, and IEEE Computer Society Digital Library. We used search terms for common data capturing modalities, such as wearable sensors, wearable devices, mobile health, patient-reported outcomes, and artificial intelligence. Years ranged from 2000 to 2023. Given the recent advancement in health information technology that is relevant to this interdisciplinary research field, we restricted the study time period to the last two decades. We searched all keywords and synonyms related to patient-generated health data, clinical decision making, health care quality, and patient safety. **Supplemental Table 1** outlines our search strategy for each database. This review was broadly inclusive of papers and focused on studies using human participants that were written in any language and available in full text (but not constrained to free article access).

### Inclusion criteria

Given the focus of this review on EHR-integrated patient-generated health data for health care applications (unlike the use of PGHD for the sole purpose of collecting data for a research protocol or without the involvement of clinicians), inclusion was that the article must have been peer-reviewed and represent empirical work (data, whether qualitative or quantitative, collected as part of the study and reported in the article). The patient-reported outcomes (PROs) were defined by the US Food and Drug Administration (FDA); PROs are “reports of the status of a patient’s health condition that comes directly from the patient, without interpretation of the patient’s response by a clinician or anyone else.”[29] PROs can be categorized as disease-related symptoms, side effects of treatment, or quality of life.[30]

### Exclusion criteria

We excluded papers that only discussed PGHD issues and case reports that focused on the architectures or infrastructures. Articles in which the data were simulated or fabricated for test purposes were also excluded. We solely included PGHD that has been integrated into EHR or health systems. The data in the studies should have been used for clinical decision support, health care quality, or patient safety. **Figure 3** provides a PRISMA flow diagram of the screening process.

**Figure 3.**
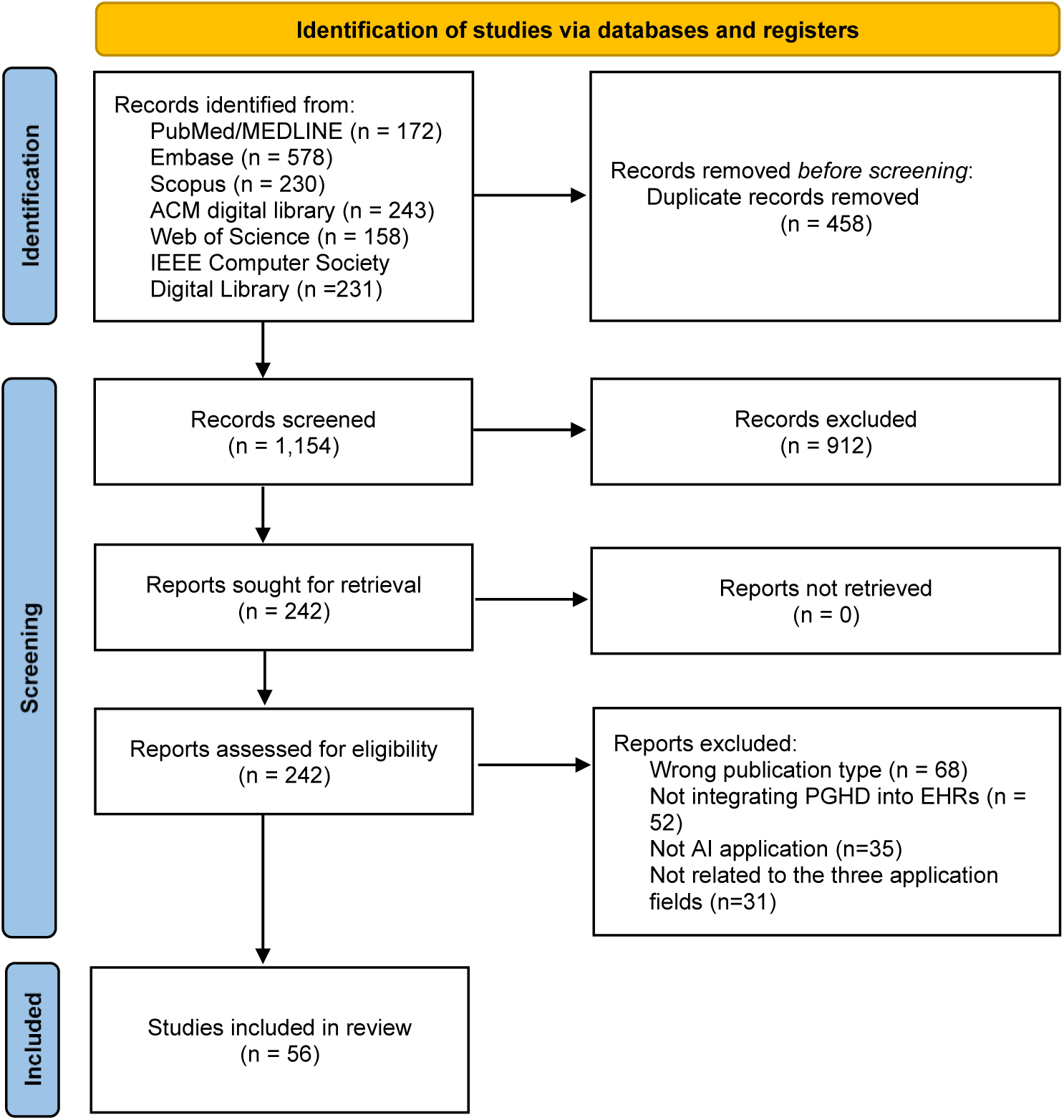
PRISMA (Preferred Reporting Items for Systematic Reviews and Meta-Analyses) flow diagram of the review process.

### Study Selection

First, duplicate articles were eliminated from the retrieved articles. Then, all the reviewers independently screened articles based on titles and abstracts to identify the studies that potentially could fit into the research question and meet the eligibility criteria. A record would be excluded if it was marked irrelevant by two reviewers. For records that were kept or difficult to decide based on the title or abstract, the full-text was scrutinized. When disagreements regarding study inclusion occurred between two reviewers, a third reviewer was involved in the discussion until consensus was reached. A total of 56 publications met the review criteria after screening.

## RESULTS

### Artificial intelligence’s capacities

AI has gained significant attention in the past decades, particularly in the context of improving health and well-being.[9] AI works by combining large amounts of data with fast and iterative processing using intelligent algorithms, allowing the system to identify and learn patterns or features from the data automatically.[31] **Table 1** demonstrates domains, theories, methods, and technologies in AI.

**Table 1.** AI theory, technologies, and their applications on EHR-integrated PGHD system.

| AI theory and technologies | Definition | Roles of AI in the applications of EHR-integrated PGHD |
| --- | --- | --- |
| **Machine learning (ML)** | ML automates analytical model building. It uses methods from mathematics, statistics, and operation science to find hidden insights in the integrated data.[91] | ML can quickly analyze large amounts of PGHD and offer actionable insights. The advanced algorithms are developed and combined in new ways to analyze more data faster and at multiple levels. This intelligent processing is key to understanding complex systems,[92] identifying and predicting target events,[93] optimizing health system workflows,[94] and improving clinical process management.[95] |
| **Deep learning (DL)** | DL leverages huge neural networks,[96] like neurons that process information by responding to external inputs and relaying information between each unit, with many layers of processing units. Leveraging the advances in computing power and improved training techniques, DL can learn complex patterns in large amounts of data. | Instead of organizing data to run through predefined equations, DL sets up basic parameters about the data and trains the machines to learn on its own by recognizing patterns using many layers of processing. Patients can use their portable device to take medical images [97] or use smartphones to record voice and send to providers.[98] All these types of PGHD could be analyzed by DL. |
| **Computer vision (CV)** | CV relies on pattern recognition and deep learning to analyze the information in the videos or pictures.[99] | Using digital images from cameras or videos and DL models, machines can accurately identify and classify objects — and then make reactions. CV can facilitate the analyses of patient-generated medical images or videos in real-time manners. |
| **Natural language processing (NLP)** | NLP enables the machine to organize, analyze, understand interpret, and manipulate human languages.[100] | NLP makes it possible for machines to read text, hear speech, interpret it, measure sentiment, determine which parts are important, and identify the root cause of human dissatisfaction.[101, 102] NLP can improve the communication between humans and computers by allowing natural language interaction.[103] |

### Integration of EHR and PGHD

EHR data include health information of clinical checkups, medical procedures, and drug prescriptions, and is focused on individual patients. In a traditional health care environment, providers typically make decisions based on the data they collect in clinical care settings. These data create a snapshot of the patient’s health at single points in time rather than continuous measurements outside of clinical settings.[4] Traditionally, clinicians can only leverage EHR data to develop diagnosis and treatment plans. EHR can be combined with other high-value datasets, such as administrative and claims data, genomic data, and other personal health information to develop rich longitudinal profiles of individual patients and populations.[32] Analytic opportunities regarding PGHD are envisioned in the context of big data and AI in medicine. AI applications are more effective when they can integrate large amounts of data, such as PGHD and EHR, both of which present diverse facets of health. PGHD has been envisioned to supplement EHR data, with a rich picture of a person’s environmental context, behavioral patterns, and lifestyle information.[33] Filing in the clinical visit’s gaps using PGHD has the potential to inform better clinical decision making, with patients’ active engagement in the process.[34]

Integrating PGHD into EHR adds value by expanding the scope of biomedical, public health, and population health research that can be conducted, which would not have been feasible by relying on EHR data alone. EHR-integrated PGHD can capture the patient’s voices and perspectives, amplify real-life situations, and strengthen the patient-provider relationship, thus improving clinical decision support, health care quality, and patient safety.

### Clinical decision support

The EHR-integrated PGHD can empower patients to manage their health and collaborate with clinicians to improve the quality of clinical decision-making. Clinicians can gain a better understanding of the patient’s health over time and reduce unnecessary visits or hospital admissions, and patients can undergo targeted interventions in the preoperative setting to mitigate the massive burden on the health care system postoperatively, all of which could contribute to better patient satisfaction and save medical resources.[34] Patient reported outcome, as one example of PGHD, is particularly important when patients need to make a decision from multiple treatment plans that demonstrate similar benefits regarding survival. PRO data can inform decision-making in this situation by providing insight into quality of life, such as the ability to maintain one’s roles and responsibilities during treatment and the ability to engage in physical activity, which can also have significant financial benefits.[35] In addition, PROs may be less vulnerable than traditional clinical assessments to “white coat syndrome”, in which patients are less likely to report symptoms during an interview.[36] **Table 2** demonstrates the roles of AI in the application of integrated EHRs and PGHD in clinical decision support.

**Table 2.** Roles AI in the application of the integrated EHRs and PGHD in clinical decision support.

| Domains | Definition | Roles and functions of AI-assisted EHR-integrated PGHD system | Examples |
| --- | --- | --- | --- |
| Administrative function/ automation | Diagnostic code selection, automated documentation and note auto-fill. | Facilitate the scheduled review and approaches for acquiring and implementing new clinical knowledge.<br>Simplify the data entry and documentation workflow. | Epic Systems Corporation (Verona, WI) is a common EHR that has a library of PROs and allows patients to add their own. Via a patient portal, users can trigger PROs at particular set time points or in response to a particular clinical event. [104] |
| Clinical management | Adherence to clinical guidelines, follow-up and treatment reminders. | Help clinicians and care teams gain a more holistic view of their patients' health and understand how contributing factors may influence health outcomes and quality of life.<br>Improve the interactions between clinicians and patients by aligning with patients' health needs and goals, ultimately increasing patient engagement and adherence to care plans and improving care delivery.<br>Provide more holistic views of a patient's health and quality of life over time, which increases visibility into a patient's adherence to a treatment plan or study protocol and enables timely intervention before a costly care episode. | Researchers have trained algorithms to analyze images of retinas taken using a smartphone-based device and diagnose this disease with over 90% accuracy.[105] |
| Diagnostics support | Providing diagnostic suggestions based on patient data and automating output from test results. | Increase trust by providing direction for patients to update their knowledge in case they were not aware of the recommendation.<br>Provide equitable opportunities for patients with limited mental and communication capabilities to record and share information about their health with their caregivers.<br>Provide tailored support, advocating on behalf of the patient, and engaging in discussions about care coordination and the patient's care plan. | AI can deploy technologies like image recognition, NLP, and deep learning to quickly detect life-threatening conditions and assess risks for diseases like injury,[106] brain cancer [107] or heart disease.[108] |
| Patient decision support | Decision support administered directly to patients through PGHD system. | • Empower patients and caregivers to manage their health by collaborating with clinicians via shared decision-making that considers patients' preferences.<br>• Enable patients to participate in the data collection process, observe how their health may fluctuate over time, and understand how certain actions and behaviors may influence their health outcomes. | Using advanced machine learning, AI has also been leveraged to analyze EHR-integrated PGHD with a diverse array of datasets and identify potential biomarkers that indicate the onset of deterioration ranging from a concussion to coma.[109] |
| Cost containment | Reducing test and order duplication, suggesting cheaper medication or treatment options, automating tedious steps to reduce provider workload. | • Gain a better understanding of the patient's health by receiving and analyzing PGHD between clinical encounters.<br>• Periodic review of data can reduce unnecessary clinician's office for routine, in-person visits, and travel costs.<br>• Ensure that patients remain in good health by dynamically monitoring the integrated data and avoid costly escalations in care, such as emergency room visits.<br>• Monitor patient data between clinical visits to intervene timely to prevent hospital visits or other costly care encounters. | Through NLP and other pattern recognition tools, machines can rapidly process EHR-integrated PGHD and automatically transcribe clinical notes. Automation can free up time and reduce costs by eliminating manual data entry. |
| Workflow improvement | Improving and expediting an existing clinical workflow with better retrieval and presentation of the integrated data. | • Facilitate the establishment of the workflow and "chain of command" that help to reduce organizational and clinician liability concerns by assigning accountability and responsibility through clearly defined procedures.<br>• The electronic processes reduce the amount of effort required by providers and reduce the potential human errors. | AI algorithms draw upon EHR-integrated PGHD to better identify patterns and assist clinicians in making diagnoses and developing treatment plans. AI can also help doctors diagnose diabetic retinopathy, one of the world's leading causes of blindness, by using image recognition.[105] |

AI plays a vital role in leveraging the integrated data and creating prediction models of outcomes, including complications, readmissions, and mortality risk. Advanced statistical algorithms and modeling, including descriptive modeling (to dissect patterns and relationships in the observed data), predictive analytics (to predict future events based on existing data), and statistical hypothesis testing (to assess causal inference), can enable a comprehensive and rigorous analysis of the integrated heterogeneous data sets. We acknowledge that AI’s goal is not to replace physicians’ clinical judgment but to help them rapidly prioritize patients’ symptoms and assess a range of diagnostic possibilities.

### Health care quality

EHR-integrated PGHD has contributed to improvements in quality of care, fewer readmissions, less resource utilization, and increased consumer satisfaction.[37] A study has shown that integrated data facilitated a better patient-provider relationship, which contributed to more efficient modes of accessibility of health care and an increased sense of empowerment of patients.[38] **Table 3** demonstrates the roles of AI in the application of integrated EHRs and PGHD in health care quality. We mapped the findings on the STEEEP (safety, timeliness, effectiveness, efficiency, equitable, and patient-centeredness) framework.[39]

**Table 3.** Roles of AI in the application of the integrated EHRs and PGHD in health care quality.

| STEEEP Domains | Definition | Roles and functions of AI-assisted EHR-integrated PGHD system application | Examples |
| --- | --- | --- | --- |
| Safe | Avoid harm to patients from the care that is intended to help them. | Analyze large volumes of PGHD, including free-text and unstructured data and supplement them with claims data and other health-related data to monitor adverse events and predict and track the spread of infectious diseases earlier with greater accuracy than traditional prediction methods. | AI helps reduce administrative burdens and burnout by correcting human errors intelligently.[63, 110] Identifying drug to drug interactions or overdose because different medications were prescribed with the same chemical drug and clinically inappropriate scheduling.[49, 111] |
| Timely | Reduce waiting time and delays for both those who receive and those who give care. | Clinicians gain a better understanding of the patient's health over time and reduce office visits and hospital readmissions.<br>Periodic reports from the AI-driven system can make it less necessary for healthy patients to spend time in the clinician's office for routine, in-person visits.<br>By having access to these data, clinicians can intervene sooner, ideally when potentially negative indicators are first observed, and can make changes to a patient's treatment protocol or instruct the patients or their caregivers timely. | Health data such as heart rate, blood sugar, and blood pressure have been collected passively using remote monitoring technologies without clinical visits;[112, 113] patients can fill out PRO surveys either via a portal, on the internet, or via a smartphone application that connects with the EHR system.[104] |
| Effective | Provide services based on scientific knowledge to all who could benefit and refrain from providing services to those not likely to benefit (avoiding underuse and misuse, respectively). | Patients may be more prepared for clinical visits because they have captured and reviewed their data, received the clinical instructions from the AI-driven system, and may have identified their own concerns and discussion points before the visit. | PGHD has been validated to improve survival rate by triggering an email alert to a clinical nurse,[114] which was a landmark event in oncology and was noted by the American Society of Clinical Oncology in their online major milestones.[115] |
| Efficient | Avoid waste, including equipment, supplies, ideas, and energy. | With the PGHD, the care team does not have to capture this data during a patient encounter and can use their time more efficiently, potentially enabling them to see more.<br>Optimize the pre-visit and check-in, which eliminates wait times at clinicians' offices, improves patients' ability to communicate with the | Sage Bionetworks launched mPower as a study using surveys and sensors to track symptoms of Parkinson's disease.[116] The study showed that the integrated data could help patients, caregivers, and clinicians better understand changes in |
|  |  | clinical staff, and enables efficient registration in multiple EHR systems. | disease over time and the impact of physical activity or medication. |
| Equitable | Provide care that does not vary in quality because of personal characteristics such as gender, ethnicity, geographic location, and socioeconomic status. | • Provide accessibility for patients who live far from clinics or may have disabilities or other serious conditions that impede travel. The availability of these data enables telehealth options, such as virtual visits, to become an alternative to in-person visits by using remote monitoring to minimize the need for clinician capture of vital signs and status updates.<br>• By using tools that generate, collect, and share PGHD, increased opportunities exist for patients to receive equitable health care service and deliveries. | Verily Health's Onduo program, for instance, embedded a mobile application into a smart device and offered virtual care for people with type 2 diabetes thereby standardizing the data and treatment recommendation regardless of racial and gender characteristics thereby reducing bias.[117] The smart device could measure blood glucose levels. It also provided information on nutrition and medication management. The app also offered a coaching dimension that identified lifestyle patterns and provided patients feedback to improve their health. |
| Patient-centered | Provide care that is respectful of and responsive to individual patient preferences, needs, and values and ensuring that patient values guide all clinical decisions. | • Track and analyze patients' health status that was expressed by their own language over time and generate data reports with visualizations that facilitate their understanding of their health.<br>• The health care can be personalized to patients' needs and lifestyles by analyzing patients' health goals, habits, preferences, and priorities.<br>• With the use of PGHD, shared decision-making between patients and clinicians is improved, and patient understanding of and adherence to treatment plans rises. | The work done in transplant surgery using integrated data reduced readmissions from 58% to 28% at 90 days.[118] Virtual health care, coupled with data from patient-reported surveys, wearable sensors, smartphone apps, digital therapeutic devices, at-home diagnostics, and social networking services, could be used to make better clinical decisions and to support better patient outcomes.[119] |

Levering AI algorithms, the integrated data can help clinicians develop personalized health care recommendations for patients. PGHD that are generated through wearable devices are unique because they are continuously, passively, and objectively collected in free-living conditions; such data are different from those generated through traditional technologies that require the manual input of data.[40, 41] AI-assisted, EHR-integrated PGHD systems can improve and increase access to treatment for populations in resource-limited settings.[42] For example, there is growing evidence that AI-driven chatbots, through voice recognition and assistance, can address routine patient questions and help clinicians send messages about diagnosis results, risk evaluation, and treatment plans to patients.[43]

### Patient safety

In most health care computerized systems, a health care worker (e.g., nurse) is needed at the interface between the data capture and patient visits. As the modes of data interpretation progress, AI is providing more sophisticated algorithms to interact with patients in an automated and efficient way and to better predict outcomes.[44] For example, computer vision facilitated the interpretation of images and videos by machines at or above human-level capabilities including object and scene recognition.[45] Augmented reality (AR) and virtual reality (VR) can be incorporated at every stage of a health care system, which can make users focus on the task without ever being distracted by moving their visual fields away from the region of interest.[46] This is a great opportunity to develop better systems to capture and interpret the EHR-integrated PGHD. Some PRO data collection apps, such as Carevive (Carevive Systems Inc), have used AI algorithms to suggest self-management strategies.[47] Some studies have also shown well-articulated, measure-specific algorithms for identifying severe symptoms and clinically significant worsening of symptoms. [48] **Table 4** demonstrates the roles of AI in the application of integrated EHRs and PGHD in patient safety. We mapped the findings on the WHO global patient safety framework.[49]

**Table 4.** Roles of AI in the application of the integrated EHRs and PGHD in patient safety.

| WHO patient safety framework | Definition | Roles and functions of AI-assisted EHR-integrated PGHD system | Examples |
| --- | --- | --- | --- |
| Eliminating avoidable harm | Make zero avoidable harm to patients a state of mind and a rule of engagement in the planning and delivery of health care everywhere. | Automatically monitor patient safety performance of the organization.<br>Providing reliable real-time data, agreed metrics, benchmarking and best practices to inform evidence-based planning. | Utilization of PGHD may take place via a web-based application on the patient's smartphone, handheld tablet, or home computer.[120] The care team can review the data and trigger a particular response, which is an opportunity to prevent, intervene, or mitigate symptomatology or impending complications.[121] |
| High-reliability systems | Build high-reliability health systems and health organizations that protect patients from harm. | Provide rapid recommendations from analyses of adverse events and through proactive risk management.<br>Promote transparency with patients by ensuring that patients have access to their medical records with an interpretable format.<br>Establish a clear specification of roles and responsibilities based on the analytical system to identify, mitigate, and eliminate risks to patients and providers.<br>Widen multisectoral collaboration to support and prioritization of safety. | The All of Us research program initiated a Fitbit Bring-Your-Own-Device project, which allowed participants to connect their Fitbit account to share data with researchers, including physiological data such as heart rate, physical activity, and behavior data like sleep. [122] These studies underscore the potential secondary uses of PGHD for generating health insights from a large real-world population that might not have been possible using conventional methods of data collection. |
| Safety of clinical processes | Assure the safety of every clinical process. | Systematically identify the sources of risk and harm in the whole process of health care, and to formulate patient safety solutions for different settings and share their expertise.<br>Monitor medication-related harm by integrating with the existing pharmacovigilance system.<br>Perform routine, regular surveillance of health care-associated risks to guide interventions with rapid feedback of results to health workers, stakeholders and public health authorities. | Group Health created an electronic Health Risk Assessment (e-HRA) on its MyGroupHealth portal to gather health risk and health history information from patients. The clinical team reviewed the patient data and integrated it into electronic health records (EHR).[123] With the integrated data, the profile functions could deliver immediate prompts to the care teams. |
|  |  | Strengthen early and accurate pathogen identification and antimicrobial resistance results to guide the most effective and safest patient treatment using the right drugs, doses, and duration of treatment.<br>Provide support in adapting and implementing patient safety strategies and interventions across the care continuum, including home, primary care, and transitions of care. |  |
| Patient and family engagement | Engage and empower patients and families to help and support the journey to safer health care. | Provide mechanisms for patients and families to co-design safer health care processes.<br>Optimize the care processes and reorient them to make services patient-centered.<br>Provide a platform for non-discrimination, patient autonomy, informed consent and shared decision-making, emergency response, access to medical records and full disclosure of adverse events.<br>Create a patient safety reporting mechanism that encourages patients and families to report and, by collecting, collating, and analyzing patient-reported experiences and outcomes of unsafe care, demonstrate actions for learning and improvement. | As the face-to-face time of traditional clinical visits is always limited, the value of PGHD lies in enriching the interaction between the health care provider and patients to facilitate shared-decision making.[124] This bidirectional dialogue contributes to patient satisfaction and a sense of autonomy or agency. |
| Health worker education, skills, and safety | Inspire, educate, skill and protect health workers to contribute to the design and delivery of safe care systems. | Facilitate the appropriate and fair duration of deployments, working hours, rest breaks, as well as minimizing the administrative burden on health care workers to prevent burnout and enhance overall well-being.<br>Assess system's acuity and could also support principles of highly reliable organizations (HROs). | In the US, with the enactment of the American Recovery and Reinvestment Act of 2009 (ARRA),[125] there is a push for widespread adoption of health information technology (HIT) through the Health Information Technology for Economic and Clinical Health Act (HITECH).[126] Hospitals and clinics are integrating EHRs and computerized physician order entry (CPOE) within their health information systems. |
| Information, research and | Ensure a constant flow of information and | Identify patient safety priorities that need to be addressed immediately. | Kaiser Permanente used secure messaging to enable patients to ask questions, inquire about |
| risk management | knowledge to drive the mitigation of risk, a reduction in levels of avoidable harm, and improvements in the safety of care. | Establish or adjust the reporting and learning system to an appropriate scale according to the capacity of the organization to capture, analyze, and investigate incidents. | test results, communicate concerns, seek clarification, and report on adverse effects.[127]<br>The study found that secure messaging with clinician review decreased the amount of clinical visits, increased measurable quality outcomes, and improved patient satisfaction. |
| Synergy, partnership and solidarity | Develop and sustain multisectoral and multinational synergy, partnership and solidarity to improve patient safety and quality of care | Provide opportunities for inter-organizational collaborative initiatives and setting up schemes to allow the organization's health care workers to exchange problem solving and improvement ideas across different systems and settings. | PGHD has introduced additional benefits during the COVID-19 pandemic. Because of social distancing and quarantine policies, health care providers have to get closely acquainted with virtual platforms such as telehealth.[128, 129]<br>By leveraging PGHD, virtual health care delivery and services can improve accessibility and effectiveness through transmission of real-time clinical data.[130] |

The integration of PGHD and EHR is another method of assessing recovery and expectations.[50] Studies have reported some successful alerts and feedback mechanisms based on the EHR-integrated PGHD, including: out of range vital signs; [51] patient alerts to contact clinicians if experiencing a symptom severity of grade 3 or higher; [52] symptom-treatment mismatch notice to the patient with alerts sent to a clinician; [53] dashboard showing asthma control with notification of local air pollution levels as well as medication adherence; [54] alert on a dashboard for nurses if individual blood pressure measurements cross a specified critical level;[55] a patient app and provider dashboard, including the ability to graph findings over time;[56] feedback about the comparison between entered blood glucose value and patient-specific target; [57] alerts for symptoms that pass critical thresholds with feedback about what and when to report to the providers;[58] and dashboard flagged patients with specified alerts, such as isolation or persistent symptoms, based on movement and communication or if patient’s response indicates thoughts of self-harm.[59]

Diagnostic errors are another major problem in the healthcare system and patient safety, with most patients experiencing at least one diagnostic error in their lifetime.[60] AI promises to help physicians accurately diagnose medical conditions in their patients and treat disease at an early stage. By deploying machine learning applications and other AI methodologies, integration of the EHR-integrated PGHD flows into a single descriptive and predictive data analysis and modeling framework that can generate better algorithms for care.[61]

### Challenges

While leveraging AI in the application of the integrated EHRs and PGHD promises to benefit patients and clinicians, challenges must be overcome to realize that potential. The large volume of data requires stakeholders to determine and invest in data storage and technical architecture to support the analysis, which can be used to serve clinical use and workflow redesign.[62] In addition, to review the PGHD, health care providers may have to allocate time from the actual patient visit that is not remunerated separately.[63]

Given the variations in accuracy, usability, and validity, some PGHD may not yet be fit for clinical use where data quality is paramount.[64] The quality of the data collected by a home monitoring device may be insufficient.[65] For example, home monitoring of heart failure patients has been broadly used, but the outcome has not been improved.[45] The lack of efficacy may be fed in part by excessive reliance on remote monitoring of weights and symptoms, which are likely late markers of incipient heart failure decompensation.

Undeveloped interoperability standards are one of the challenges of PGHD. Some systems develop their own technical infrastructure for PGHD.[66] The lack of standards that fully address PGHD use cases and consensus on which interoperability standards to use lead to variations in data representation and coding that limit the exchange, normalization, and completeness of the integrated data. This hinders the system’s ability to draw valuable insights. The lack of standardized terminology and format for PGHD also limits secondary uses of the data in research studies and clinical trials.[67]

Ensuring the security and privacy of PGHD is also a challenge. EHR-integrated PGHD may be at risk for security breaches that could affect data integrity and expose the data to access for malicious purposes because they may not subject to the same security regulatory framework as HIPAA-regulated entities.[68] Concerns include insecure points of data collection and movement that potentially expose the device or the clinician’s information system to pollutants. In addition, there is growing potential for risks related to unauthorized access, including cyber threats.

### Recommendations

Using AI to clean and filter the PGHD before integration is helpful to reduce the volume of the data streams. To prevent the duplication of records when integrating PGHD with EHR, clinicians and researchers can employ AI-assisted patient matching techniques. Current procedures that use statistical algorithms to match data in local EHR systems with PGHD have shown increased levels of reliability. Technologies like the application programming interfaces (APIs) are portable packages of code that make it possible to add AI functionality to existing software packages and products.[69] They can add capabilities for image recognition to home security systems, calling out patterns and insights of interest in PGHD. APIs could also facilitate data accuracy, privacy, and security by their authentication function. The Office of the National Coordinator for Health Information Technology (ONC) is leading a PCOR Trust Fund project on Patient Matching, Aggregating, and Linking that aims to use APIs to enable linking of patient data, including PGHD, to other clinical and claims data.[70] AI tools are capable of cleaning integrated data and simplifying the process of ensuring data are complete and prepared for analysis. These electronic processes reduce the volume of data and amount of effort required to clean and manage the integrated system and reduce the potential for human errors.

User verification solutions, such as biometric authentication and multi-step identity verification, could increase data accuracy and validity. Graphical processing units (GPU) provide outstanding computing power for iterative processing,[71] which enables the integration of PGHD and EHR, health information exchange (HIE), and higher speed calculation. Big data companies enable the use of predictive analytics such as natural language processing (NLP) and machine learning (ML) on structured or unstructured data, coupled with GPUs, to help doctors and hospitals make their data more usable.[72] Data brokers, gateways, and aggregators could assess and manage data lineage and accuracy.[73] Research has also shown that the accuracy of AI applications and models could be developed with greater accuracy if the algorithms were to be shared publicly.[74]

The emerging data exchange standard, Fast Healthcare Interoperability Resources (FHIR), and APIs provide opportunities to flexibly create software that securely pulls discrete data from the EHR into third-party software.[75] FHIR could be used to improve the presentation and visualization of the integrated data. FHIR’s latest extensions, SMART Markers and SMART-on-FHIR, are also approaches developed to streamline and simplify data integration.[75] The interface needs to have digital flexibility so that data exchange is convenient and versatile.[76] Leveraging standards-based data exchange through interoperability could potentially solve both the interoperability challenges as well as ease PGHD integration,[77] making it an achievable goal for more health care systems. [78]

Blockchain is a list of records that are linked together using cryptography, which is a distributed ledger—write once and never erase.[79] Blockchain promotes health data management,[80] provenance, exchange, and sharing.[81] It can also be leveraged to improve the privacy and security of EHR-integrated PGHD.[82] This emerging technology creates a distributed, digital ledger of cryptographically secure transactions and provides solutions that support security, privacy, and interoperability between EHRs and IoT devices while enabling trust in the validity of the data and its source.[83] Direct Secure Messaging, which was developed in 2010 under ONC’s Direct Project, could achieve security, privacy, data integrity, and user authentication during the exchange of health information over the Internet.[78]

## DISCUSSION

PGHD provides the potential for a holistic perspective of people’s health conditions by presenting rich information about their lives. PGHD captured by digital health and informatics tools, such as wearable devices, portable point-of-care devices, smartphone apps, PROs through online questionnaires, and connected IoTs, can allow patients to become more engaged in the process of receiving health care.[84] The AI-driven application of the EHR-integrated PGHD has the potential to close some health care gaps caused by the limitation of EHRs and support personalized medicine, including empowering patients and families to engage via shared decision-making, improving patient-provider relationships, and reducing the time and costs of clinical visits. Challenges mainly stem from the large volume of integrated data, data standards, data exchange and interoperability, security and privacy, interpretation, and meaningful use. In addition, the use of PGHD must be implemented in a way that prevents the exacerbation of health disparities.[85] Pernicious disparities, such as gender or race differences in health conditions related to social inequality within health data, have negative impacts on AI model performance and results.[86] Developers of such software should draw heavily from research in the fields of user-focused design and human-computer interaction to create intuitive data visualizations that focus on important clinical information and allow health care providers to identify patterns that provide clinically meaningful insights.

The use of PGHD in health care is at a promising stage and inevitable but needs further work for widespread adoption and seamless integration into health care systems. AI has the capacity to optimize PGHD integration into EHRs considering resources, standards for data exchange, security and privacy, and clinical workflows. With the EHR-integrated PGHD, AI can help transform health care by improving diagnosis, treatment, and the delivery of clinical care. AI-based computing systems can greatly improve clinicians’ abilities to diagnose their patients’ health issues, classify risks at a patient level by drawing on the power of the integrated data, and provide much-needed support to clinics and hospitals in under-resourced areas.[87] AI can also expand the operational capacity of EHR-integrated PGHD systems and streamline manual tasks in the health care system to boost productivity.[88]

EHR-integrated PGHD allows for transformation from historically aggregated, population-based data to individual, longitudinal data, and more granular data, where more advanced methodologies need to be applied to identify patterns, changes, and outliers.[89] AI provides advanced methodologies for interpreting EHR-integrated PGHD, including predictive analytics (which uses various techniques to predict future events based on existing large data sets), machine learning (which uses computer systems to complete tasks relying on inferences over time), and deep learning (which focuses on learning data representations), and other complex analyses. PGHD allows patients, caregivers, and families become part of the integrated health care system by incorporating richer information from time and space dimensions.[90]

## CONCLUSION

The use of PGHD in health care is at a promising stage but needs further work for widespread adoption and seamless integration into health care systems. AI-driven, EHR-integrated PGHD systems can greatly improve the capacity to deal with the large volume of data, control data quality, enable data exchange with better interoperability, ensure security and privacy, make understandable interpretations, and generate more meaningful use for health care. With EHR-integrated PGHD, AI can help transform health care by improving clinical decision support, health care quality, and patient safety. Further efforts are needed to understand how EHR-integrated PGHD systems can better support health care with the assistance of AI.

## Data Availability

This is a review article.

## FUNDING

None.

## CONTRIBUTION STATEMENT

J.Y. designed the study, developed the review protocol and search strategies, screened the references, extracted the data, assessed the quality of the included references, and contributed to the analyses. All the authors were involved in the screening process of the references, extracted the data, and the writing of the manuscript. All the authors read and approved the final version of the manuscript.

## CONFLICT OF INTEREST STATEMENT

None.

## ACKNOWLEDGEMENT

The authors would like to thank Linda O’Dwyer for her help with the development of the search strategies.

## Supplemental Tables

**Supplemental Table 1.**
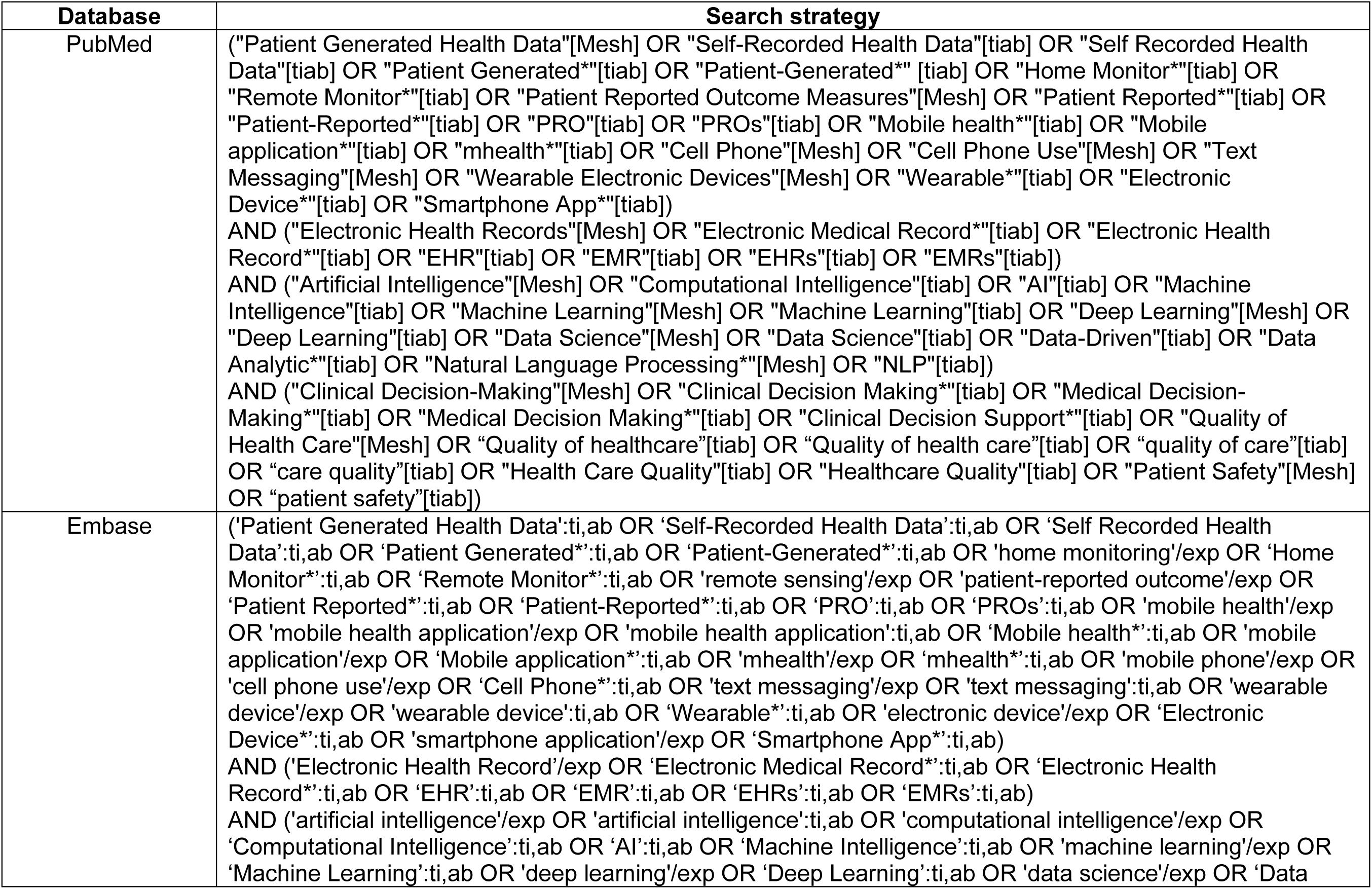
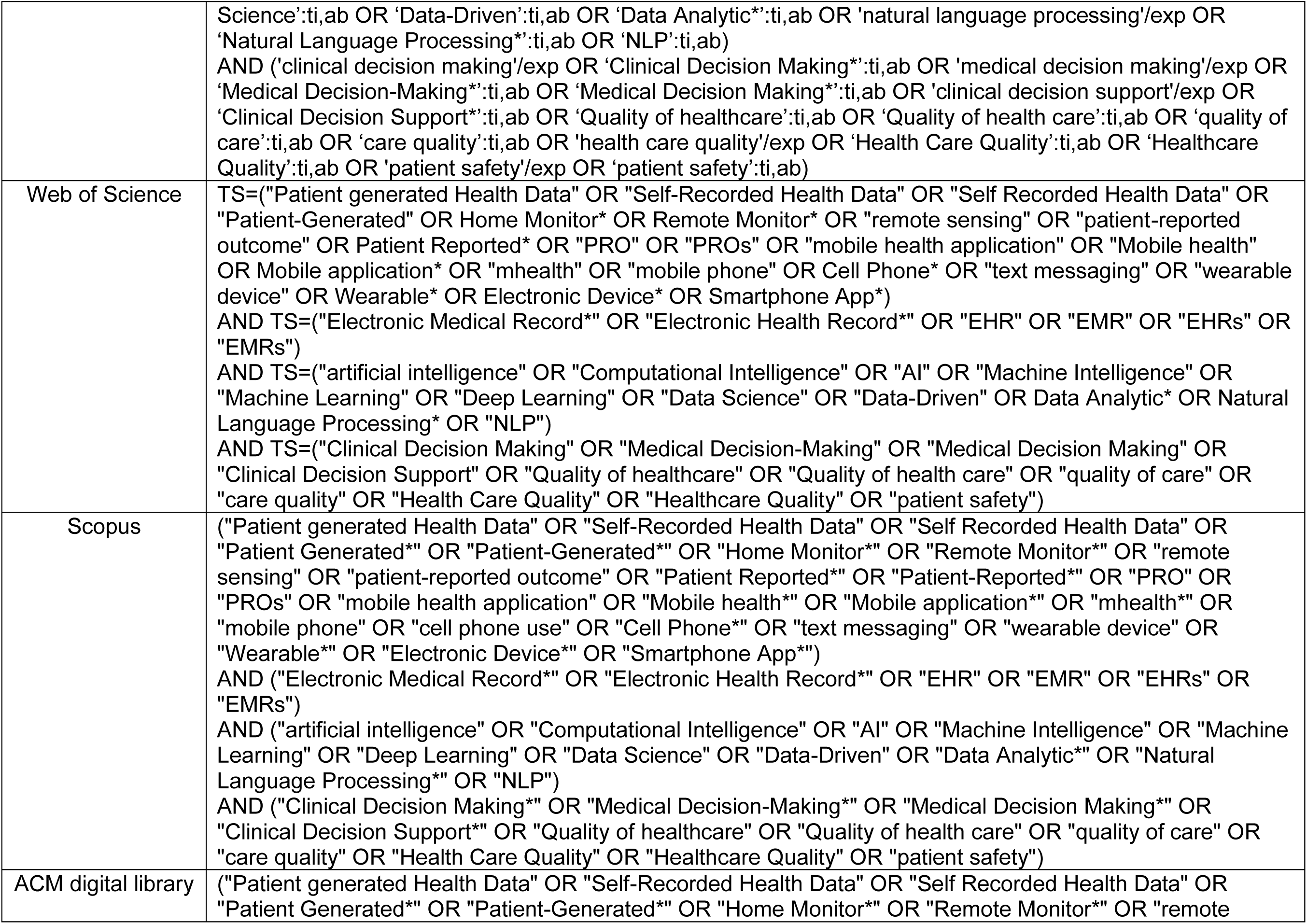
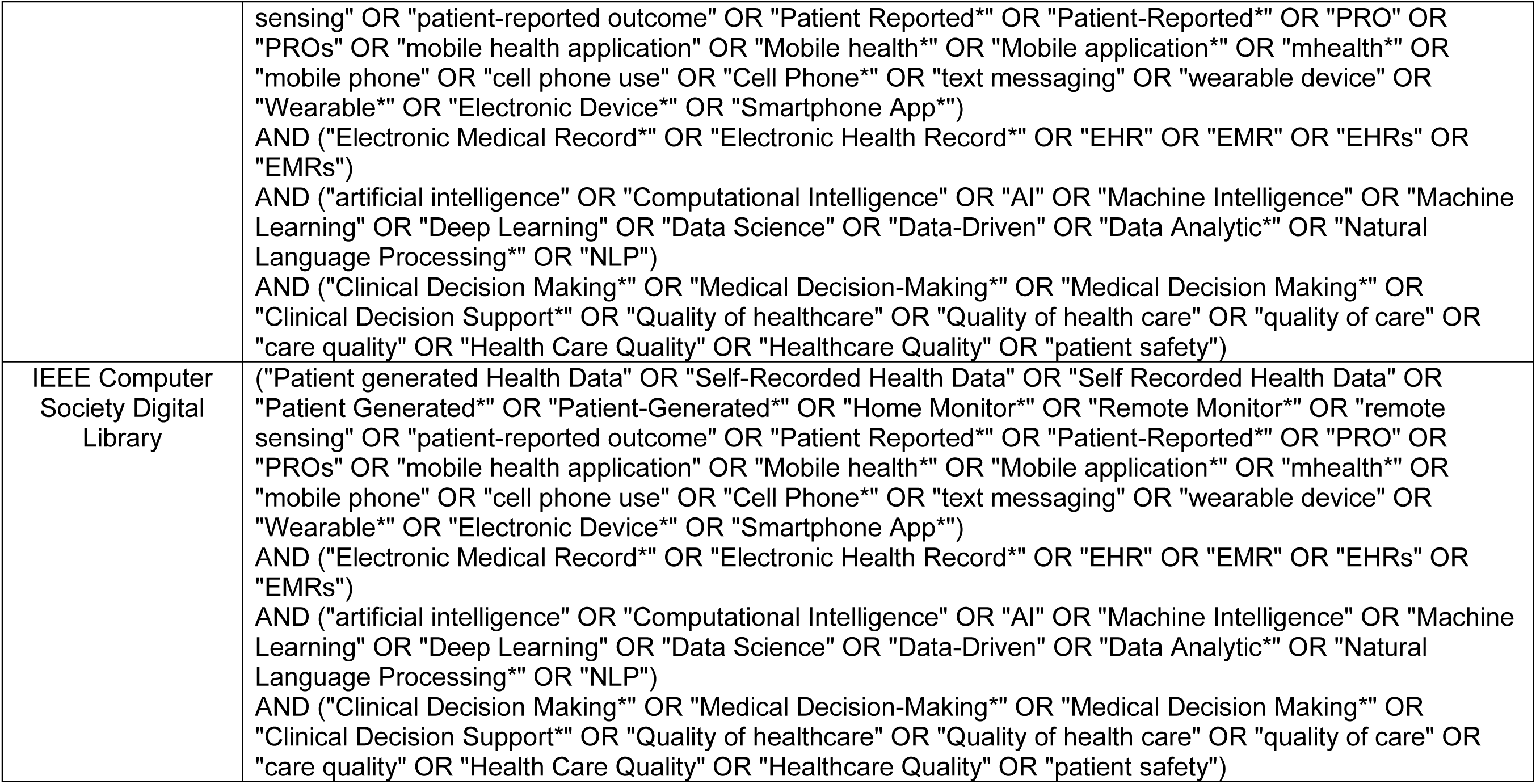
Search Strategy Outline.

## Notes

### Competing Interest Statement

The authors have declared no competing interest.

### Funding Statement

No funding.

### Summary of Updates

Author affiliations updated in the paper

